# Decoding Restenosis Risk After CABG: A Combined Transcriptomic and Mendelian Randomization Analysis

**DOI:** 10.1101/2025.08.06.25333136

**Authors:** Muhammad Ainul Yaqin, Fadhiil Ansyarullah Murtadho

## Abstract

**Background:** Restenosis following coronary artery bypass grafting (CABG), especially in saphenous vein grafts (SVGs), remains a significant long-term complication, affecting up to 20% of cases. While technical and biological contributors are well characterized, non-technical lifestyle and clinical risk factors remain underexplored due to limitations in observational research. This study aimed to identify non-technical risk factors contributing to restenosis after CABG by integrating transcriptomic data and Mendelian randomization analysis.

**Methods:** We analyzed microarray data (GSE241205) to identify differentially expressed genes (DEGs) between occluded and unoccluded SVGs. Enrichment analysis via DAVID identified key biological pathways. The most enriched pathway—immune system activation—was used as the outcome in two-sample Mendelian randomization using GWAS summary statistics for 15 lifestyle and metabolic exposures. Causal estimates were calculated using inverse variance weighted (IVW) method, with sensitivity analyses for pleiotropy and heterogeneity.

**Results:** Among 4,443 DEGs, IGKV3D-20 (log2FC 14.02) and OTC (log2FC -7.29) were the most significantly altered. Immune system activation, particularly complement activation, emerged as the dominant pathway. Mendelian randomization revealed high LDL (>159 mg/dL; OR = 12.57), total cholesterol (>200 mg/dL; OR = 11.20), smoking (16 cigarettes/day; OR = 9.98), and alcohol intake (250 ml/day; OR = 3.24) significantly increased pathway activation. Protective factors included physical exercise (30 mins/day; OR = 0.22), HDL (<40 mg/dL; OR = 0.41), aspirin (OR = 0.83), and clopidogrel (OR = 0.24). No significant pleiotropy or heterogeneity was observed.

**Conclusion:** Immune activation plays a central role in restenosis after CABG. Modifiable non-technical factors—particularly lipid profile and lifestyle behaviors—contribute significantly and may guide preventive strategies.

## INTRODUCTION

Coronary artery bypass grafting (CABG) is a prevalent procedure for treating coronary artery disease (CAD) and continues to be the most frequently performed cardiac surgery[1, 2]. Although alternative treatments such as percutaneous coronary intervention (PCI) are gaining popularity, CABG remains the gold standard for revascularization in patients with multivascular CAD, especially those with severe blockages[3–5]. The effective conduit options are the saphenous vein, the internal mammary arteries, the radial artery, and the right gastroepiploic artery[6]. Saphenous vein grafts (SVGs) are the most often utilized conduits for CABG. nonetheless,

Overall, postoperative restenosis occurs in around 20% of vein grafts[7–10]. SVGs were associated with a high incidence of restenosis in 10 years[11]. The early restenosis occurring in the initial weeks after the CABG is primarily caused by technical factors and thrombotic events. On the other hand, restenosis after the first month of CABG is mainly attributed to neointimal hyperplasia, which subsequently leads to the development of atherosclerosis and becomes the primary cause of late restenosis[12]. Neointimal hyperplasia is driven by systemic conditions (such as metabolic diseases, atherosclerosis, and hypertension) and local factors (such as adaptation to left-sided circulation)[13–16]. Information related to biological mechanisms and operative technical factors has been widely explored, but the identification of risk factors related to lifestyle to prevent restenosis after CABG has not been comprehensively accommodated by the literatures.

Several limitations have been cited as reasons for the lack of literature reporting information on non-technical risk factors for restenosis after CABG. The main limitation that has been declared is the lack of generalizability of the results due to the difficulty in obtaining restenosis models from the same population[17]. Observational studies are difficult to cover this limitation for numerous reasons including patient follow-up problems until restenosis occurs in cohort study designs and the challenge of substantial study bias in other study designs. Observational studies have three primary limitations: reverse causation, confounding variables, and bias due to sample size[18]. Studies related to potential reverse causation and interference of confounding variables in exploring risk factors for restenosis after CABG have not been conducted at all, therefore observational studies are very risky to perform because of the challenging issues of bias.

Genome-wide Association Study (GWAS) is an approach to genetic research that identifies genomic variants associated with risk of certain diseases or traits[19, 20]. The growth of GWAS produces many summary statistics that encourages researchers to use these data to identify risk factors for a disease, including restenosis after CABG. The use of secondary data from GWAS is called Mendelian Randomization Analysis, a statistical methodology that defines exposure through genetic instrumental variables and outcome through genes associated with certain biological pathways[21, 22]. This technique accommodates the issue of bias previously declared against observational studies. The limitation of time-consuming and ineffective study design can also be tolerated by Mendelian randomization analysis[23]. However, there has been no association of restenosis after CABG with specific genes or biological pathways. Therefore, we identified genes and biological pathways that play a role in restenosis after CABG and used them as outcomes to identify non-technical risk factors involved in the development of these pathways. We identified relevant genes and biological pathways using integrated bioinformatics analysis and analyzed their risk factors using Mendelian randomization analysis.

## METHODS

We performed continuous and integrated bioinformatics analysis using microarray data of transcriptome sequences from occluded vein grafts after CABG. **Figure 1** shows the flow of our method. We used publicly available data generated in accordance with the guidelines in the Helsinki Declaration. Hence, ethical approval was not applicable.

**Figure 1.**
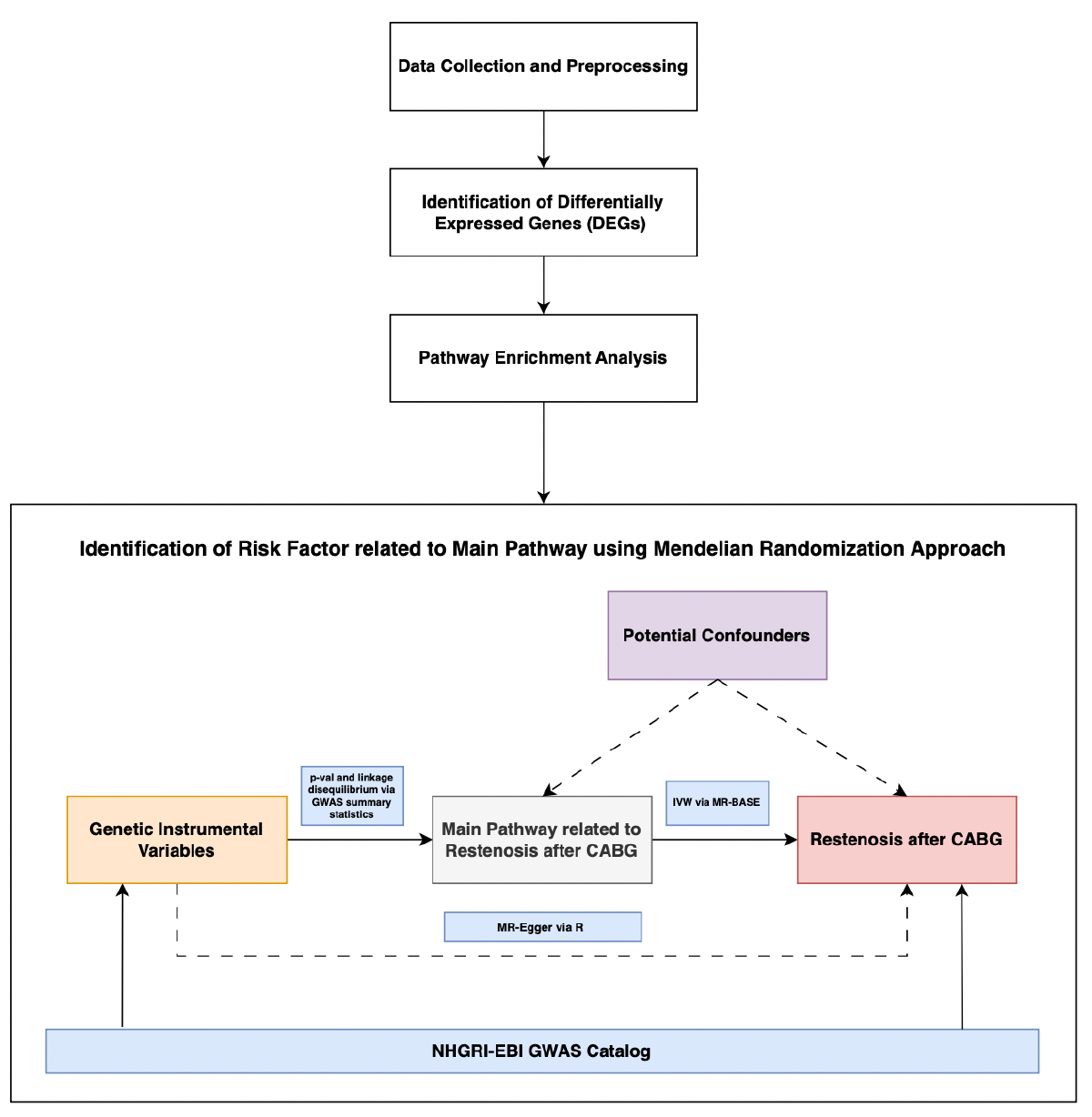
Flow of Integrated Bioinformatics Analysis with Mendelian Approach

### Data Collection

NCBI-GEO (https://www.ncbi.nlm.nih.gov/geo/) is a functional genomics database that stores transcriptomic sequence data internationally. From this database, we retrieved one gene dataset from transcriptomic sequencing of occluded vein grafts after CABG, i.e. GSE241205 provided by Liu et al.[24]. The Gene Dataset includes three occluded vein graft samples that underwent clinical redo-CABG, three un-occluded samples, and 58,939 genes.

### Identification of Differentially Expressed Genes (DEGs)

The DEG between the occluded and unoccluded sample groups was identified using the GEO2R tool with statistical significance set to |log2FC| > 0 and P value < 0.05. We downloaded the normalized data in the form of FPKM (Fragments Per Kilobase per Million mapped) value for exploratory data analysis. Then, we selected 10 upregulated genes with the highest log2FC and 10 downregulated genes with the lowest log2FC to represent the DEGs data in the subsequent analysis. Volcano plots, correlograms, and heat maps of the 10 upregulated and downregulated genes were generated using the web-based application SR Plot (https://www.bioinformatics.com.cn/en).

### Pathway Enrichment Analysis

Gene ontology (GO), which consists of three separate ontologies (cellular component, molecular function, and biological process), is the most extensive and commonly utilized knowledge base for gene functions. The Kyoto Encyclopedia of Genes and Genomes (KEGG) is a database resource for learning about biological systems. Our study uses the Database for Annotation, Visualization and Integrated Discovery or DAVID (https://david.ncifcrf.gov/) to identify the DEGs’ functions and pathway enrichment. Then, we used the SR Plot (https://www.bioinformatics.com.cn/en) to visualize the GO and KEGG terms of the DEGs. The P value < 0.05 was the cut-off criterion. This stage aims to discover the possible pathways DEG takes in its contribution to restenosis after CABG. Our risk factor analysis selected the majority pathway with the highest enrichment score as the represented variable of outcome.

### Risk Factor Analysis

Mendelian Randomization Analysis is a method to infer causality between exposure and outcome using genetic proxies. We performed a two-sample unidirectional Mendelian randomization analysis. In our study, to define risk factors, we used different genetic proxies obtained from the collaboration of National Human Genome Research Institute and European Bioinformatics Institutes (NGHRI-EBI) GWAS Catalog (https://www.ebi.ac.uk/gwas/). We curated robustly significant Single Nucleotide Polymorphism data associated with 15 risk factors with p < 5×10^−8^ and r^2^ > 0.0001. The profile of each risk factor and its genetic proxy is shown in **Supplementary Table 1**.

For potential pathways obtained through enrichment analysis in the previous stage, we selected genetic instruments through the MR Base Web Application (https://www.mrbase.org/). Using the database mining tools of MR Base, we conducted Mendelian randomization analyses to estimate the effect of each factor on the risk of pathway development. For primary Mendelian randomization analyses, we combined per-SNP effects using inverse variance weighted (IVW). Reported estimates were converted to odds ratios where the outcome was binary and interpreted using a conservative p-value threshold (0.05/number of factors with available summary statistics). A horizontal pleiotropy test was carried out using MR-Egger where p-intercept > 0.05 is the cutoff for no significant pleiotropy. To detect heterogeneity, we also used MR-Egger for obtained Q-statistics and their p-value > 0.05 is the criteria for no significant heterogeneity.

## RESULTS

### Genomic Exploratory Data Analysis and Identification of DEGs

We retrieved the raw expression data of each gene from the GSE241205 dataset consisting of three unoccluded vein graft samples (GSM7719383, GSM7719384, and GSM7719385) and three occluded vein graft samples (GSM7719386, GSM7719387, and GSM7719388). The correlation between the six samples used in this study is shown in **Figure 2A**. The Pearson correlation results indicated that the sample quality was reliable.

**Figure 2.**
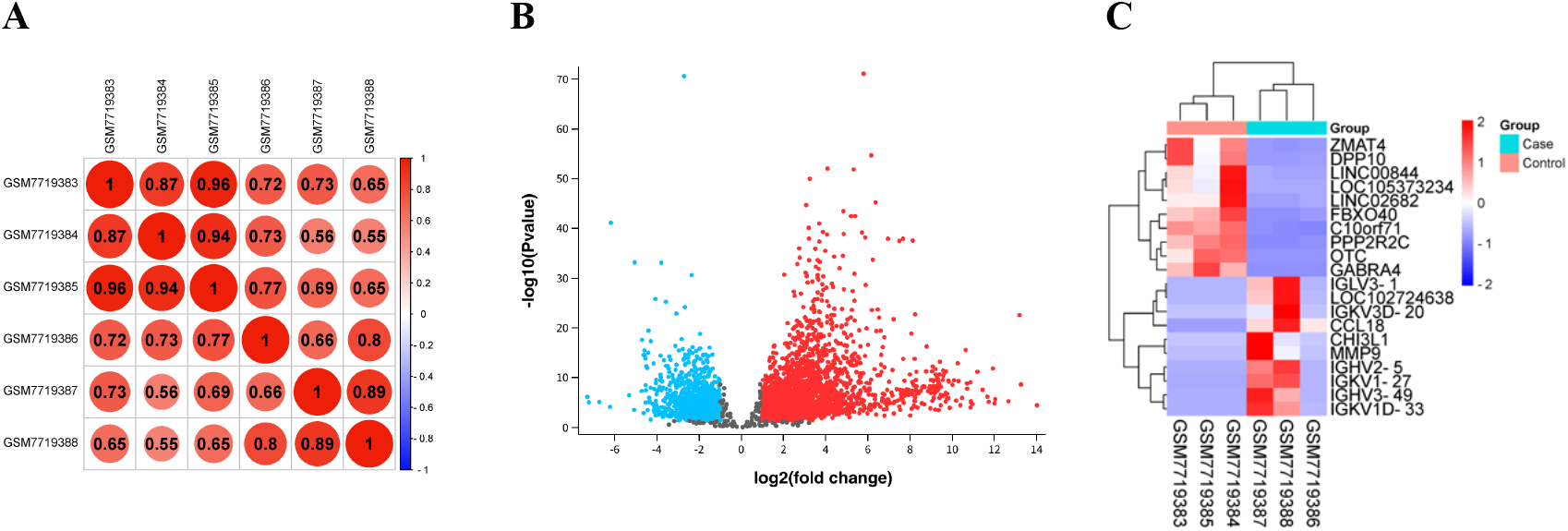
Functional Genomic Exploratory Data Analysis. Correlogram of six gene gxpression samples (A); Volcano Plots of 4,443 differentially expressed genes (B), Heatmap of top 10 upregulated and downregulated differentially expressed genes (C)

From the two groups of sample datasets, 4,443 DEGs were obtained with 1,594 downregulated and 2,849 upregulated. A volcano plot of these DEGs is shown in Figure 2B. The upregulated DEGs with the most robustly different expression between the occluded vein grafts and unoccluded vein grafts sample groups were IGKV3D-20 (Immunoglobulin Kappa Variable 3D-20) with log2FC 14.02 and p = 3.8 × 10^−6.^ The downregulated DEGs that showed the most difference in expression among the sample groups was OTC (Ornithine Transcarbamylase) with log2FC -7.29 and p = 5.17 × 10^−8^. **Figure 2C** shows the heatmap of the top 10 upregulated and downregulated DEGs.

### Pathway Enrichment Analysis

GO and KEGG analysis was performed on the top 10 upregulated and downregulated DEGs to identify the biological role of these genes and their pathways to cause restenosis after CABG. **Figure 3A** shows the Gene Ontology terms for each cluster i.e. biological process (BP), cellular component (CC) and Molecular Function (MF). GO analysis of the selected DEGs by DAVID showed significant enrichment in the component associated with immunoglobulin complex with robustly affected restenosis by the classical pathway of the complement activation process. In addition, the enriched MF included antigen binding, immunoglobulin receptor binding, etc. **Figure 3B** accommodated the results of pathway enrichment analysis by KEGG terms. The top DEGs significantly contributed to the restenosis after CABG through immune system activation, especially in complement activation and humoral immunity.

**Figure 3.**
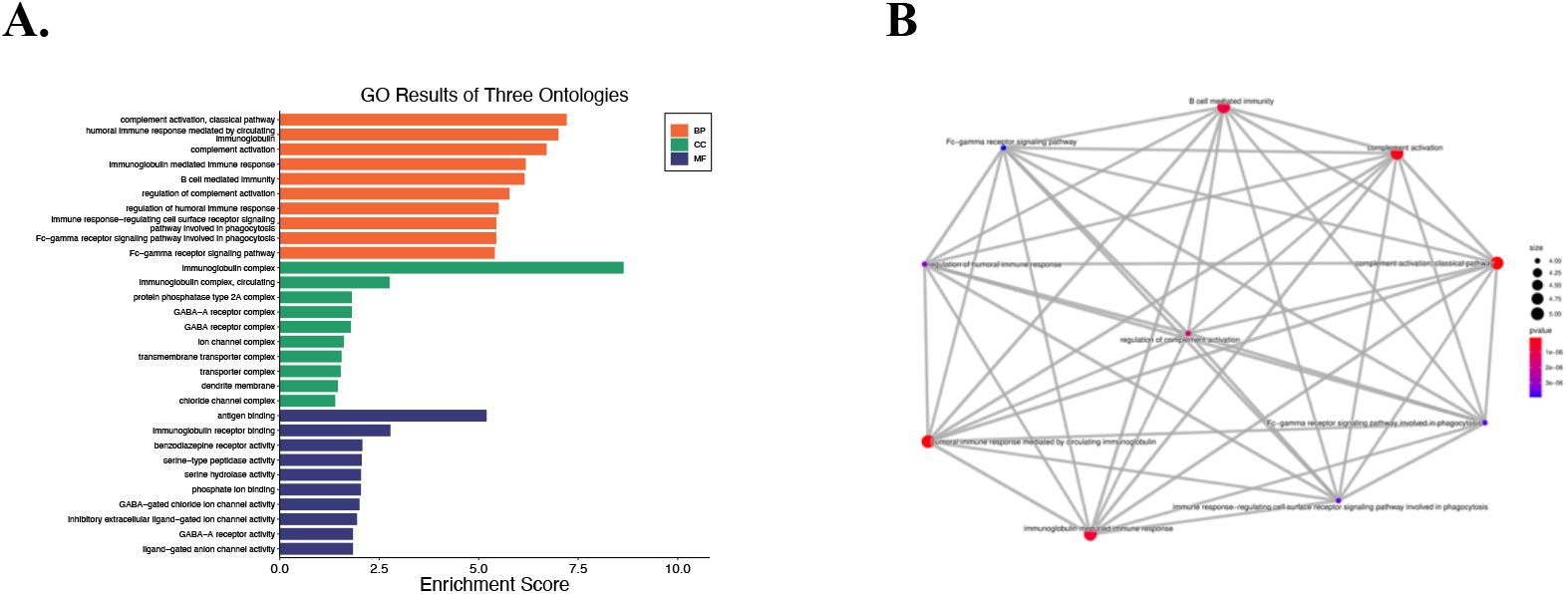
Pathway Enrichment Analysis. Gene Ontology Terms (A); Kyoto Encyclopedia of Genes and Genomes Network Terms (B)

### Risk Factor Analysis

We tested several risk factors in the connectivity with immune system activation as a determinant pathway for restenosis after CABG using available GWAS summary statistics through Mendelian randomization analysis. Unidirectional Mendelian randomization analyses between each risk factor and immune system activation revealed several findings suggesting causal relationships in the IVW methods frameworks. **Figure 4** arbitrates the connectivity findings using the IVW method. From the analysis, three risk factors that were not significant in affecting restenosis-related immune system activation were tea consumption, high triglyceride levels, and high diastolic blood pressure. In addition, we identified eight factors that increased the risk of developing a pathway for restenosis after CABG. Conversely, four risk factors reduced the risk of developing restenosis-related biological pathways.

**Figure 4.**
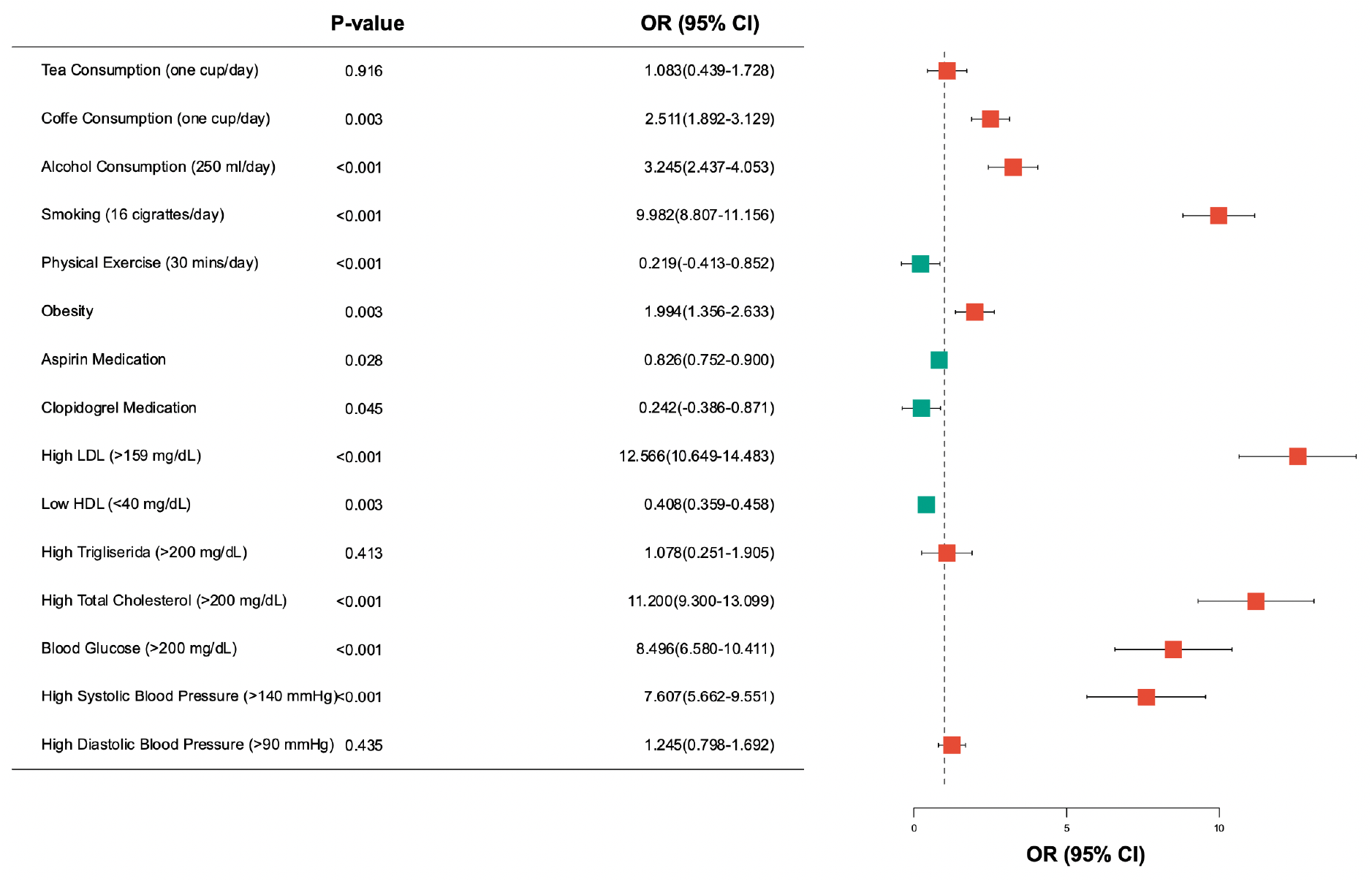
Risk Factor of Restenosis-related Pathway

LDL with levels greater than 159 mg/dL was the risk factor that most positively contributed to pathway activation associated with restenosis after CABG (OR = 12.57; 95%CI = 10.65 - 14.48; p < 0.001). Physical exercise for 30 minutes per day was the factor that significantly reduced pathway activation (OR = 0.22; 95%CI = -0.41 - 0.85; p < 0.001). In line with LDL, total cholesterol levels of more than 200 mg/dL also increased the risk of restenosis after CABG through the immune system activation pathway (OR = 11.20; 95%CI = 9.30 - 13.10; p < 0.001). Lifestyle and smoking habits with 16 cigarettes/day (OR = 9.98; 95%CI = 8.81 - 11.16; p < 0.001), coffee consumption of one cup/day (OR = 2.51; 95%CI = 1.89 - 3.13; p = 0.003), and alcohol consumption of 250 ml/day (OR = 3.24; 95%CI = 2.44 - 4.05; p < 0.001) also positively contributed to the development of the restenosis pathway. Other protective factors that reduced the restenosis-related pathway were aspirin medication (OR = 0.83; 95%CI = 0.75 - 0.90; p = 0.028), clopidogrel medication (OR = 0.24; 95%CI = -0.39 - 0.87; p = 0.04), and HDL < 40 mg/dl (OR = 0.41; 95%CI = 0.36 - 0.46; p = 0.003).

**Table 1** accommodates the possibility of pleiotropy and heterogeneity between the datasets we used in the risk factor analysis. MR-Egger showed no significant horizontal pleiotropy (p-intercept > 0.05). Using the same method, our inclusion data did not indicate any significant heterogeneity (p > 0.05).

**Table 1.**
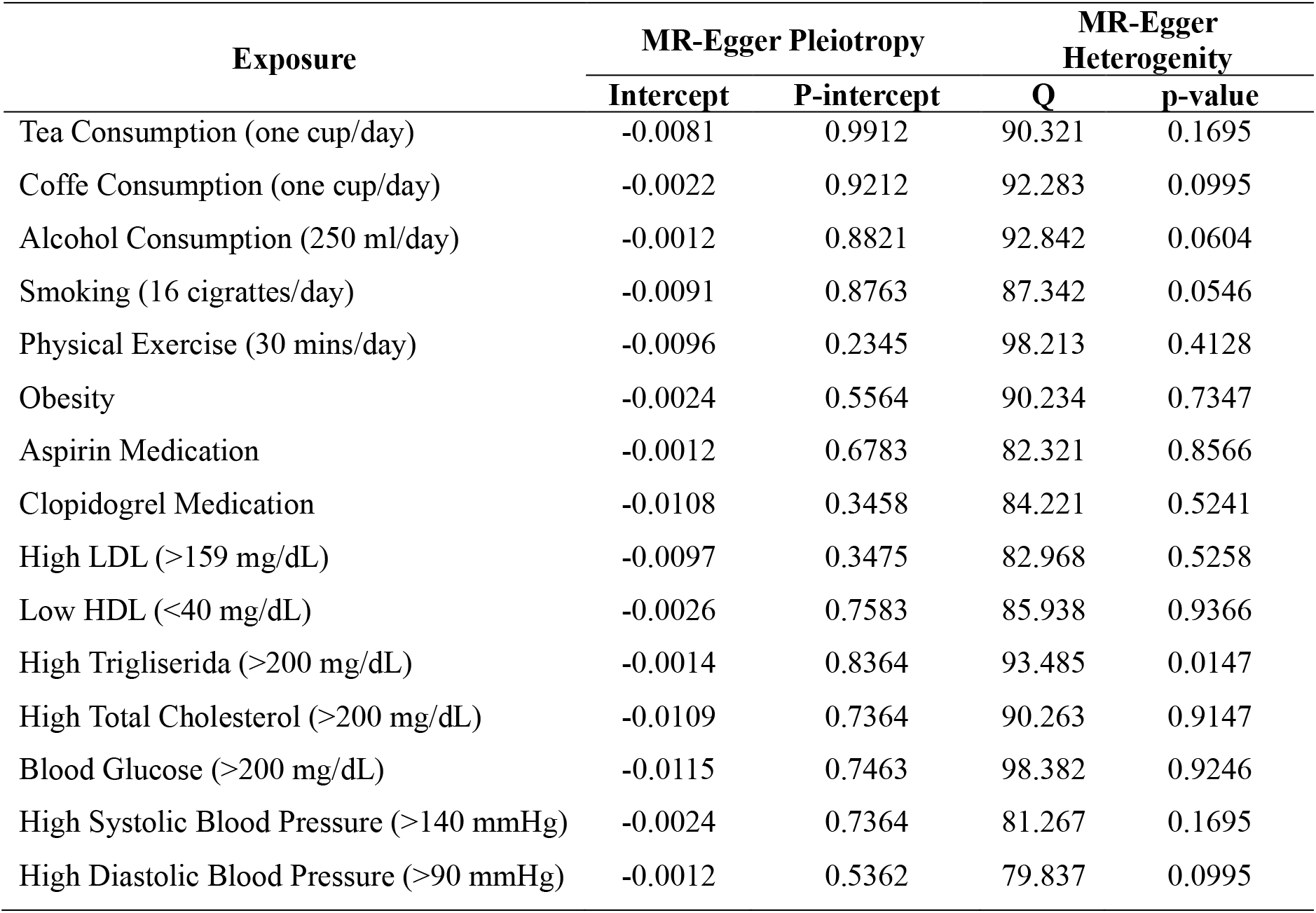
Pleiotropy and Heterogeneity between the Datasets.

| Exposure | MR-Egger Pleiotropy |  | MR-Egger Heterogeneity |  |
| --- | --- | --- | --- | --- |
|  | Intercept | P-intercept | Q | p-value |
| Tea Consumption (one cup/day) | -0.0081 | 0.9912 | 90.321 | 0.1695 |
| Coffe Consumption (one cup/day) | -0.0022 | 0.9212 | 92.283 | 0.0995 |
| Alcohol Consumption (250 ml/day) | -0.0012 | 0.8821 | 92.842 | 0.0604 |
| Smoking (16 cigrattes/day) | -0.0091 | 0.8763 | 87.342 | 0.0546 |
| Physical Exercise (30 mins/day) | -0.0096 | 0.2345 | 98.213 | 0.4128 |
| Obesity | -0.0024 | 0.5564 | 90.234 | 0.7347 |
| Aspirin Medication | -0.0012 | 0.6783 | 82.321 | 0.8566 |
| Clopidogrel Medication | -0.0108 | 0.3458 | 84.221 | 0.5241 |
| High LDL (>159 mg/dL) | -0.0097 | 0.3475 | 82.968 | 0.5258 |
| Low HDL (<40 mg/dL) | -0.0026 | 0.7583 | 85.938 | 0.9366 |
| High Trigliserida (>200 mg/dL) | -0.0014 | 0.8364 | 93.485 | 0.0147 |
| High Total Cholesterol (>200 mg/dL) | -0.0109 | 0.7364 | 90.263 | 0.9147 |
| Blood Glucose (>200 mg/dL) | -0.0115 | 0.7463 | 98.382 | 0.9246 |
| High Systolic Blood Pressure (>140 mmHg) | -0.0024 | 0.7364 | 81.267 | 0.1695 |
| High Diastolic Blood Pressure (>90 mmHg) | -0.0012 | 0.5362 | 79.837 | 0.0995 |

## DISCUSSIONS

Our study analyzed risk factors that contribute to the development of biological pathways associated with restenosis after CABG. The main challenge in this study was to identify biological pathways that play a role in restenosis and make them an outcome in the risk factor analysis. To accommodate this, we applied integrated bioinformatics analysis to find biological pathways that are meaningful in developing restenosis after CABG. We then used a Mendelian randomization approach to identify risk factors associated with the development of these biological pathways. Thus, we used the two genomic approaches to achieve our research objectives.

Bioinformatics is an analytical approach that employs mathematical, statistical, and computer tools to process and evaluate biological data, as opposed to traditional laboratory work[25]. Numerous justifications for using an integrated bioinformatics analysis method to identify potential biological pathways involved in restenosis after CABG must be declared, including that this method translates biological data in the form of microarrays into health information in a time-saving framework, efficient, and cost-effective[26].

To address the challenge of identifying risk factors that contribute to the biological pathways that develop restenosis, we justify the use of mendelian randomization analysis because of its capacity to accommodate observational studies that are not possible with validated methods such as cohort or randomized controlled trials[19, 21]. Our focus on identifying risk factors that influence restenosis-related pathways is difficult to answer by conventional observational studies.

Using these two genomic approaches, we report two main results. First, the most upregulated and downregulated DEGs were IGKV3D-20 and OTC, respectively. Based on GO and KEGG analysis, the top 10 upregulated DEGs and downregulated DEGs mediate restenosis through the immune system activation pathway. This result is in line with previous studies that reported that both DEGs are genes related to immune system activation[27, 28]. The limited literature on these two DEGs contributes to the deficiency of information regarding the role of these genes in regulating health, especially restenosis after CABG. However, we were able to suggest a novel signaling pathway that contributes to restenosis after CABG. Previously, Podemska-Jedrzejczak et al have previously reported that this restenosis is associated with activation of the Vascular Endothelial Growth Factor (VEGF) pathway[29].

Secondly, we report several risk factors that contribute to immune system activation as a bridge between the role of DEGs in developing restenosis after CABG. Our study has established that high LDL greater than 159 mg/dl can increase the risk of developing restenosis-related biological pathways up to 12-fold. These results are not surprising because the guidelines for patient management after CABG state that lifestyle control and medication should result in low LDL levels[30–32]. Numerous study also states that failure to achieve cholesterol target levels following CABG has been connected with long-term mortality[33]. Biologically, much literature has described the role of LDL in atherosclerosis[34]. Atherosclerosis is a complex process involving local immune regulation of blood vessels. Endothelial inflammation, a marker of atherosclerosis, has also been reported as a determinant of restenosis[35]. This also indicates the similarity of determinants that we stated in our first important finding that the immune system is involved in restenosis.

The main limitation of this study is the use of one dataset without collaborating it with other publicly available datasets. This is based on the limited DNA Microarray dataset that targets samples with restenosis. The use of a single dataset may bias the number of genes and expression variance that are individualized and not representative of the population. This single dataset also has an impact on the limitations in conducting further analysis after pathway enrichment analysis, i.e. protein-to-protein interaction and identification of hub genes. Hub genes are genes that are highly connected to other genes in a network and are considered important for regulating that network. This stage is important in confirming the target genes that regulate most of the pathobiology of restenosis after CABG. We also did not identify causality between gene expression as exposure and restenosis as an outcome directly using Mendelian randomization analysis. This was due to the limited GWAS summary statistics related to restenosis and the absence of the previously described hub genes.

Our findings suggest the role of regulation and expression of certain genes in the development of restenosis after CABG. In addition, we also emphasize that gene expression can be regulated through the epigenomic framework of risk factor control. We found many risk factors that play a protective or associative role in the development of biological pathways responsible for restenosis after CABG. Our study impacts the advancement of genomic information in the field of cardiovascular surgery and encourages the increase of publicly available datasets on surgeries performed. Bibliometrics of the restenosis-related literature has mostly used observational approaches, so we encourage future analyses involving the understanding of genomics and bioinformatics disciplines.

## Data Availability

All data referred to in this manuscript are publicly and freely available. The transcriptomic data used for differential gene expression and enrichment analysis were obtained from the NCBI Gene Expression Omnibus (GEO) under accession number GSE241205. Summary-level genetic association data used in the Mendelian randomization analysis were retrieved from the NHGRI-EBI GWAS Catalog and the MR-Base platform, which aggregate GWAS summary statistics from multiple studies. These datasets were openly accessible to the public before the initiation of the study and did not require registration or approval for use.

https://www.ncbi.nlm.nih.gov/geo/

https://www.ebi.ac.uk/gwas/

---

**SUPPLEMENTARY TABLE 1.**
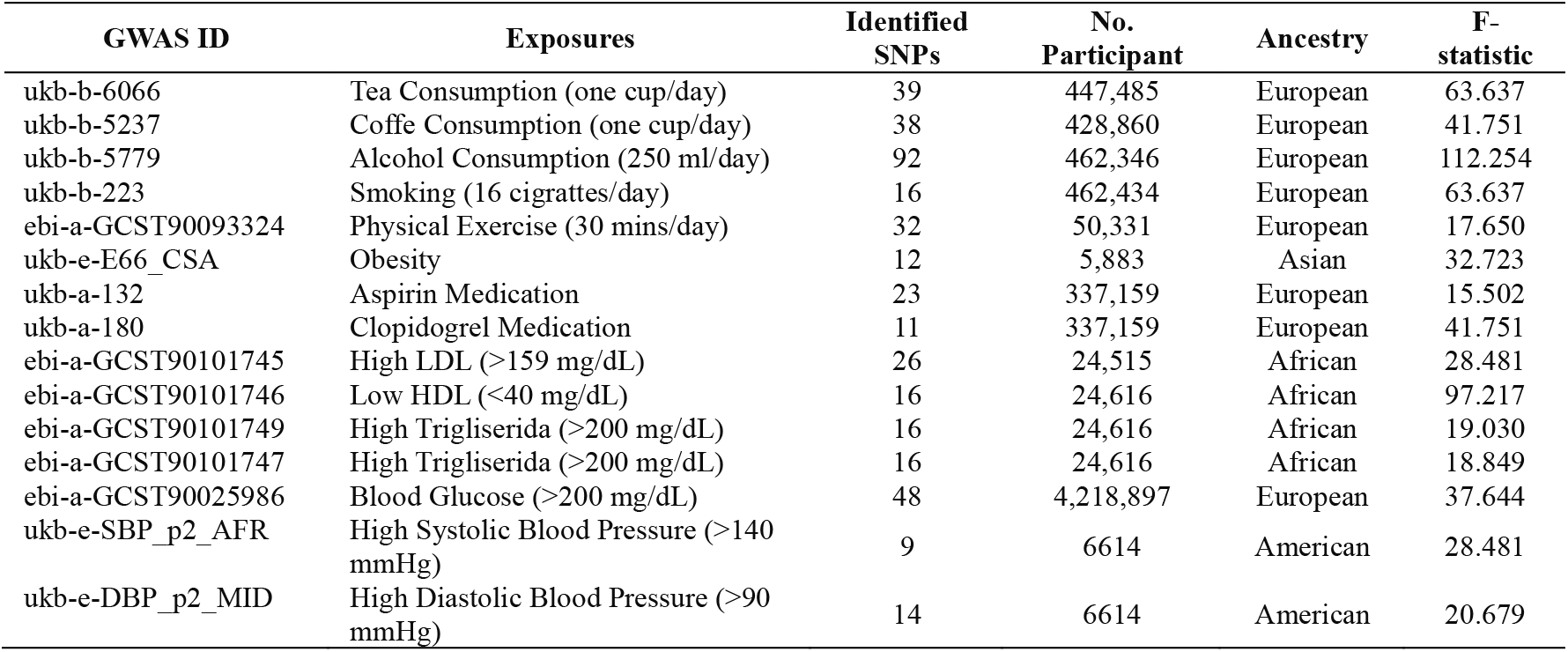

