## Supplementary Table 1 for "Decoding Restenosis Risk After CABG: A Combined Transcriptomic and Mendelian Randomization Analysis"

| GWAS ID | Exposures | Identified SNPs | No. Participant | Ancestry | F-statistic |
|---|---|---|---|---|---|
| ukb-b-6066 | Tea Consumption (one cup/day) | 39 | 447,485 | European | 63.637 |
| ukb-b-5237 | Coffe Consumption (one cup/day) | 38 | 428,860 | European | 41.751 |
| ukb-b-5779 | Alcohol Consumption (250 ml/day) | 92 | 462,346 | European | 112.254 |
| ukb-b-223 | Smoking (16 cigrattes/day) | 16 | 462,434 | European | 63.637 |
| ebi-a-GCST90093324 | Physical Exercise (30 mins/day) | 32 | 50,331 | European | 17.650 |
| ukb-e-E66_CSA | Obesity | 12 | 5,883 | Asian | 32.723 |
| ukb-a-132 | Aspirin Medication | 23 | 337,159 | European | 15.502 |
| ukb-a-180 | Clopidogrel Medication | 11 | 337,159 | European | 41.751 |
| ebi-a-GCST90101745 | High LDL (>159 mg/dL) | 26 | 24,515 | African | 28.481 |
| ebi-a-GCST90101746 | Low HDL (<40 mg/dL) | 16 | 24,616 | African | 97.217 |
| ebi-a-GCST90101749 | High Trigliserida (>200 mg/dL) | 16 | 24,616 | African | 19.030 |
| ebi-a-GCST90101747 | High Trigliserida (>200 mg/dL) | 16 | 24,616 | African | 18.849 |
| ebi-a-GCST90025986 | Blood Glucose (>200 mg/dL) | 48 | 4,218,897 | European | 37.644 |
| ukb-e-SBP_p2_AFR | High Systolic Blood Pressure (>140 mmHg) | 9 | 6614 | American | 28.481 |
| ukb-e-DBP_p2_MID | High Diastolic Blood Pressure (>90 mmHg) | 14 | 6614 | American | 20.679 |
